# The protective *PLCG2* variants delay Alzheimer’s disease onset age in *APOE ε*4 carriers

**DOI:** 10.1101/2025.09.22.25336304

**Authors:** Heli Jeskanen, Sami Heikkinen, Inka Kervinen, Jenni Lehtisalo, Tiia Ngandu, Roosa-Maria Willman, Jessica Rosa, Dorit Hoffmann, FinnGen, Ville Leinonen, Annakaisa Haapasalo, Mari Takalo, Henna Martiskainen, Mikko Hiltunen

## Abstract

**INTRODUCTION:** We investigated the impact of protective *PLCG2*-P522R and *PLCG2*-3’UTR variants, and the *TREM2*-R62H risk variant on Alzheimer’s disease (AD) onset age in relation to *APOE* ε4 status. Plasma-based biomarkers of the different variants were also explored.

**METHODS:** Kaplan-Meier and survival analyses were performed using FinnGen genotype and clinical endpoint data to assess the onset ages of AD, anxiety, and type 2 diabetes. Plasma biomarkers related to metabolism and inflammation were analyzed in 145 FINGER cohort participants.

**RESULTS:** *PLCG2*-P522R and *PLCG2*-3’UTR variants delayed AD onset, including among *APOE ε*4 carriers. *PLCG2*-P522R carriers showed elevated plasma ghrelin levels. *TREM2*-R62H variant associated with an earlier onset of AD in *APOE ε*4 carriers.

**DISCUSSION:** Protective *PLCG2* variants may mitigate *APOE ε*4-mediated risk of AD, which coincide with increased plasma levels of ghrelin. These findings highlight the further need to explore biomarkers and mechanisms associated with the protective variants in relation to *APOE ε*4.

## 1 INTRODUCTION

Genetic studies have identified a protective P522R variant in the *PLCG2* gene (*PLCG2*-P522R; rs72824905 C>G), which decreases the risk of Alzheimer’s disease (AD) and increases longevity (1–4). *PLCG2*-P522R, which potentiates PLCγ2 function, mediates several beneficial effects on microglia function in *in vivo* and *in vitro* models (5–10). Moreover, recent findings have revealed that neuronal downregulation of *PLCG2* impairs synaptic function and triggers AD-related alterations, suggesting that changes in PLCγ2 levels and subsequent functions also within neurons are significant in the context of AD (5).

PLCγ2 works downstream of triggering receptor expressed on myeloid cells 2 (TREM2) (11). TREM2 is selectively expressed in microglia and it plays a key role in modulating the cell survival, phagocytosis, and inflammatory responses (3,11,12). In *TREM2* gene, several AD associated risk increasing variants have been recognized, such as R47H and R62H (1,13–15). These variants impair TREM2 functions leading to reduced microglial activation and clearance of β-amyloid, which exacerbates the disease pathology (3,11,12). It has been suggested that when compared to non-carriers, these variants do not differ in clinical presentation of the AD at baseline but they exhibit faster cognitive decline (13).

Here, we investigated the effects of the protective *PLCG2*-P522R and *PLCG2-*3’UTR (rs4243226 A>G) variants on the onset age of AD individually and in relation to apolipoprotein E ε4 (*APOE ε*4) allele in the large FinnGen cohort originating from Finland (16). Furthermore, to identify potential biomarkers associated with the protective *PLCG2* variants, we assessed the effects of the two variants on the plasma levels of common metabolic and inflammatory markers within the well-established FINGER intervention cohort (17). Here, we show that the protective *PLCG2*-P522R variant delays AD onset age among *APOE ε*4 carriers as well as associates with increased plasma levels of ghrelin, which is known to exert anti-inflammatory effects, metabolic regulation, and cognitive enhancement. Furthermore, we show that the *PLCG2*-3’UTR variant delays the AD onset age among *APOE* ε4 carriers and in contrast, *TREM2*-R62H leads to earlier onset of AD.

## 2 RESULTS

### 2.1 Protective *PLCG2* variants exhibit minimal linkage disequilibrium

Carriership of *PLCG2*-P522R (rs72824905 C>G) showed the expected protective effect against AD in FinnGen (OR=0.55, 95% CI: 0.40-0.77, p=3.83×10^−4^). Furthermore, we identified a common variant in the 3’UTR of *PLCG2* (rs4243226 A>G, allele frequency: 0.7), which was associated with a significantly decreased risk of AD (OR=0.92, 95% CI: 0.90-0.95, p=2.55×10^−8^). Linkage disequilibrium (LD) metrics revealed r^2^ and D’ values of 0.0004 and 0.85, indicating a minimal correlation between the two variants despite the high degree of allelic co-segregation.

### 2.2 *PLCG2* variants delay the onset age of AD

To study AD onset timing and risk, we used Kaplan-Meier survival curves to illustrate differences in onset age, while the Cox proportional hazards model estimated the risk of developing AD between the genotypes. Based on the survival analyses, *PLCG2*-P522R (CG or GG) significantly delayed AD onset age as compared to non-carriers (CC) in FinnGen (Fig. 1A). Cox analysis revealed that *PLCG2*-P522R associated with a reduced risk by 42% of developing AD (HR=0.58, 95% CI: 0.42-0.82, p=0.00167). Additionally, male sex associated with an increased risk of AD (HR=1.09, 95% CI: 1.05-1.13, p=8.99×10^−7^). The *PLCG2*-P522R associated with a delayed onset of AD both in females and males (Supp. Fig. 1A-B). In the *APOE* genotype*-*stratified analysis with sex as a covariate, *PLCG2*-P522R decreased the risk of AD in the *APOE* ε3/4 group by 46% (HR=0.54, 95% CI: 0.32-0.91, p=0.022, Fig. 1B). The sex did not significantly affect the AD age of onset among the *APOE* ε3/4 carriers. However, male sex increased the risk of having AD within in *APOE* ε3/3 carriers (HR=1.20, 95% CI: 1.14-1.27, p=3.7×10^−12^, Supp. Fig. 1C-D). Within the *APOE* ε4/4 group, the *PLCG2*-P522R variant or sex did not significantly affect the AD onset age as compared to non-carrier AD patients (Fig. 1B and Supp. Fig. 1C-D). Cox regression analysis of *PLCG2*-3’UTR revealed that this common variant (AG or GG) decreased the risk of AD by 11-16% as compared to non-carriers (AA) (Fig. 1C). There were no significant differences between sexes with respect to *PLCG2*-3’UTR (Supplementary Table 1, Supp. Fig. 1E-F). According to *APOE* genotype-stratified analysis using sex as a covariate, the *PLCG2*-3’UTR variant did not significantly affect the AD onset age within the *APOE* ε3/3 group. Conversely, *PLCG2*-3’UTR moderately increased the AD onset age among homozygous *PLCG2*-3’UTR carriers as compared to non-carriers within the *APOE* ε3/4 group (HR=0.83, 95% CI: 0.76-0.91, p=4.32×10^−5^, Fig. 1D). Sex did not affect the AD onset age in the *APOE* ε3/4 group (Supp. Fig. 1G-H). Interestingly, the homozygosity of *PLCG2*-3’UTR (GG) delayed the AD onset age in the *APOE* ε4/4 carriers (HR=0.87, 95% CI: 0.78-0.96, p=0.0091, Fig. 1D). Furthermore, when sex effects were studied separately in *APOE* ε4/4 carriers, the *PLCG2*-3’UTR exerted a protective effect only in female AD patients who were either heterozygous (HR=0.74, 95% CI: 0.58-0.94, p=0.016) or homozygous (HR=0.65, 95% CI: 0.51-0.83, p=0.00051) (Supp. Fig. 1G-H) for the *PLCG2*-3’UTR variant.

**Figure 1.**
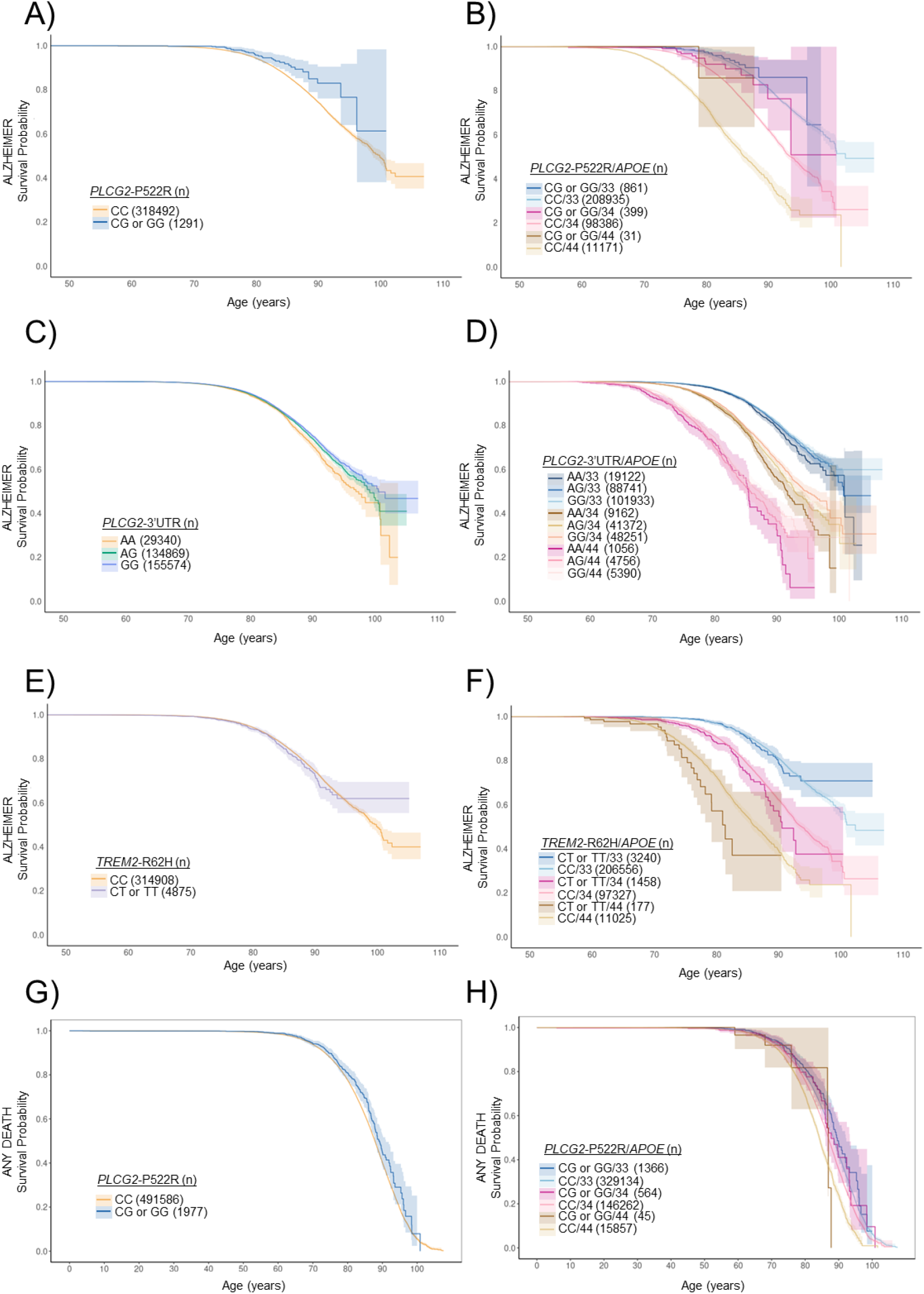
Protective *PLCG2*-P522R delays the onset age of Alzheimer’s disease. To investigate the impact of the *PLCG2*-P522R variant on AD onset, Kaplan-Meier curves on FinnGen endpoint data were utilized, with the focus on *APOE ε*3- and *APOE ε*4-carrying individuals >50 years of age. The curves illustrate the AD-free time in years, starting from age 50 until the AD diagnosis or the end of follow-up for the non-carrier group. Shaded area indicates 95% confidence intervals. A) *PLCG2*-P522R compared to non-carriers in FinnGen “ALZHEIMER” endpoint. B) *PLCG2*-P522R carriers compared to non-carriers in FinnGen “ALZHEIMER” endpoint with *APOE ε*4 allele count. C) *PLCG2*-3’UTR carriers compared to non-carriers in FinnGen “ALZHEIMER” endpoint. D) *PLCG2*-3’UTR carriers compared to non-carriers in FinnGen “ALZHEIMER” endpoint with *APOE ε*4 allele count. E) *TREM2*-R62H carriers compared to non-carriers in FinnGen “ALZHEIMER” endpoint. F) *TREM2*-R62H carriers compared to non-carriers in FinnGen “ALZHEIMER” endpoint with *APOE ε*4 allele count. G) *PLCG2*-P522R carriers compared to non-carriers in “ANY DEATH” endpoint. H) *PLCG2*-P522R carriers compared to non-carriers with *APOE ε*4 in “ANY DEATH” endpoint. *APOE*: ε3/3=33, ε3/4=34 and ε4/4=44

### 2.3 *TREM2*-R62H variant carriers show earlier onset of AD

Given that biologically PLCγ2 functions downstream of the TREM2 receptor, we also investigated the effects of the AD risk-increasing variant *TREM2*-R62H (rs143332484 C>T, OR=1.21, 95% CI: 1.04-1.40, p=1.3×10^−3^). The *TREM2*-R62H (AF=0.0078) is more common in the Finnish population than the more widely known *TREM2*-R47H (AF=0.00045) AD risk variant (15,18). No significant differences in the AD onset age were found between *TREM2*-R62H carriers (CT or TT) and non-carriers (CC) using the Cox model (Fig. 1E, Supplementary Table 2). Also, no differences were observed between females and males (Supp. Fig. 2A-B). In the *APOE* genotype-stratified analysis with sex as covariate, *TREM2*-R62H did not affect the AD onset age among the *APOE* ε3/3 carriers (Fig. 1F). Male sex increased the risk of AD to a similar extent as observed in the *PLCG2*-P522R and *APOE* ε3/3-carrying individuals (Supp. Fig. 2C-D). *TREM2*-R62H increased the risk of AD by 23% (HR=1.24, 95% CI: 1.01-1.52, p=0.04, Fig. 1F) among the *APOE* ε3/4 group. Sex did not influence the AD onset age in *APOE* ε3/4 carriers. Among the *APOE* ε4/4 carriers, *TREM2*-R62H decreased the AD onset age only in males (HR=1.80, 95% CI: 1.015-3.19, p=0.04, Supp. Fig. 2C-D).

### 2.4 *PLCG2*-P522R variant does not increase longevity but protects against anxiety

As the *PLCG2*-P522R variant has been associated with longevity (1,3), we investigated the FinnGen endpoint ‘Any Death’ to assess if the carriers lived longer than the non-carriers. However, we did not find any difference between the carriers and non-carriers (Fig. 1G). Also, there were no differences in *APOE* genotype-stratified analysis nor between sexes (Fig. 1H, Supp. Fig. 2E-H).

Previously, some *PLCG2* hypermorphic mutations have been reported to associate with an increased risk of autoimmune diseases (19). Thus, we examined whether homozygous *PLCG2*-P522R carriers exhibited signs of autoimmune diseases but found no significant associations. This observation highlights the beneficial nature of the *PLCG2*-P522R variant as opposed to strong hypermorphic mutations in *PLCG2* (20,21). In our recent study, we observed an anxiety phenotype in mice harboring the *PLCG2*-P522R (9). Accordingly, we investigated if an association with anxiety can be detected in FinnGen in the individuals carrying the protective *PLCG2*-P522R. Unexpectedly, *PLCG2*-P522R delayed the onset age of anxiety as compared to non-carriers (Fig. 2A). Notably, this effect was observed in female but not in male *PLCG2*-P522R carriers (Fig. 2A-C).

**Figure 2.**
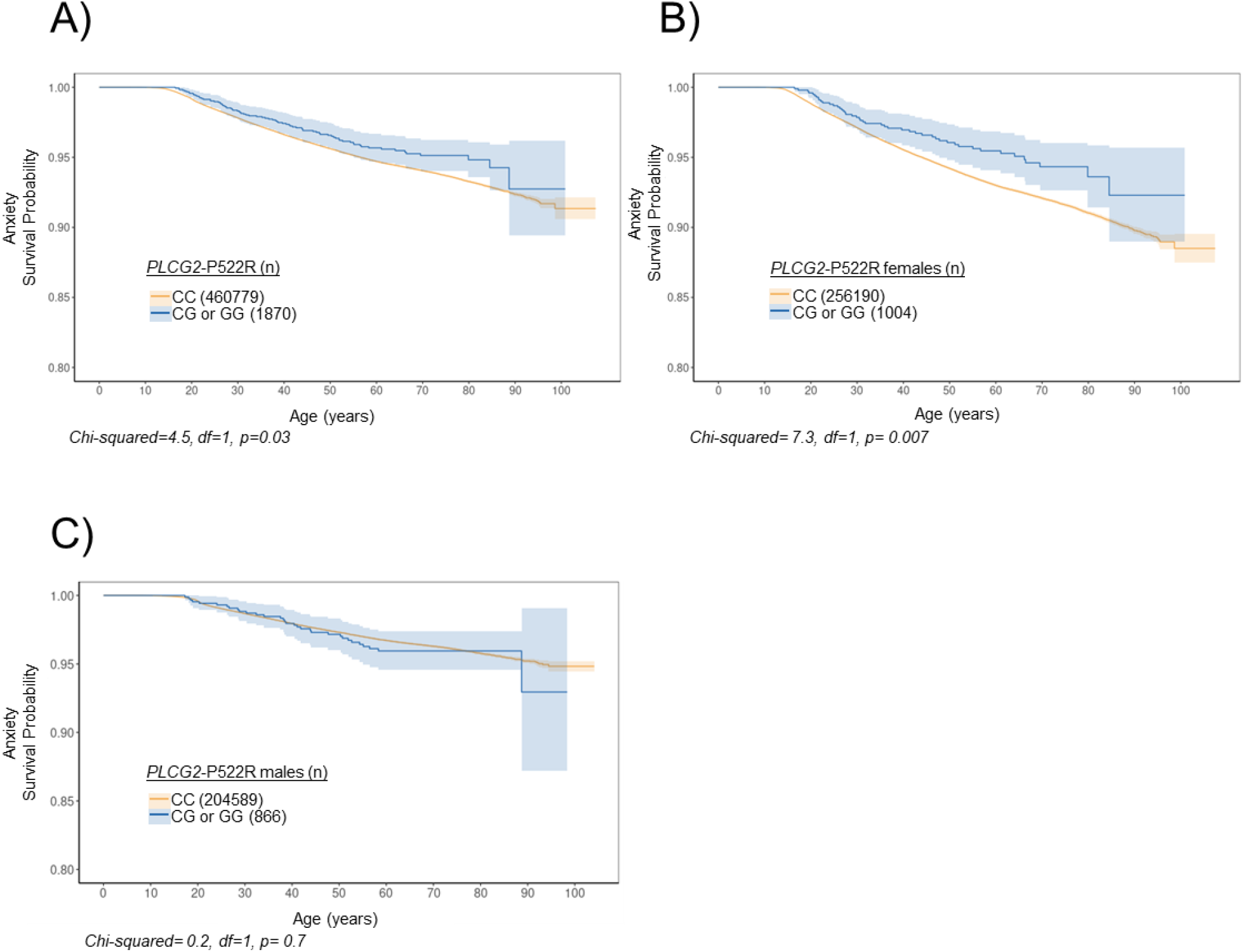
*PLCG2*-P522R is protective against anxiety. To investigate the impact of the PLCG2-P522R variant on anxiety onset, Kaplan-Meier curves on FinnGen endpoint data were utilized. The curves illustrate the anxiety-free time in years. The x-axis indicates the age at the first diagnosis for cases and the age at the end of follow-up for the non-carrier group. Shaded area indicates 95% confidence interval. A) All carriers and non-carriers, B) only females, and C) only males in “ANXIETY” endpoint.

### 2.5 *PLCG2*-P522R variant carriers have increased plasma levels of ghrelin

To identify potential biomarkers associated with the *PLCG2* and *TREM2* variants, we analyzed 40 metabolism and inflammation-related markers in plasma samples of 145 FINGER cohort individuals (17,22,23). Significantly higher levels of ghrelin and visfatin were detected in *PLCG2*-P522R carriers as compared to non-carriers or carriers of *PLCG2*-3’UTR or *TREM2*-R62H variants (Fig. 3A-B). Since ghrelin and leptin jointly regulate energy metabolism, we also examined plasma leptin levels (24). No significant differences were observed in the leptin levels between *PLCG2*-P522R and non-carriers (Fig. 3C). *APOE* genotype nor sex affected the levels of ghrelin or visfatin (Supp. Fig. 3A-E). However, males showed lower leptin levels than females (Supp. Fig. 3F). When ghrelin levels were tested using multilinear regression model adjusted by age, sex, and *APOE* genotype, the *PLCG2*-P522R was the only statistically significant effector (Supplementary Table 3). In a similar analysis, there were no statistically significant differences in the levels of visfatin or leptin (Supplementary Table 4-5). Due to the fact that ghrelin is known to increase appetite and affect peripheral metabolism (25), we investigated the waist circumference, BMI, and waist-to-height ratio of the *PLCG2* and *TREM2* variant carriers. No significant differences in these parameters between the genotypes were observed (Fig. 3D-F). Furthermore, levels of plasma C-reactive protein (CRP), an indicator of inflammation, remained unaffected at baseline and during seven-year follow-up measurements between the carriers and controls (Supp Fig. 4). Interestingly, when screening metabolism-associated endpoints, *PLCG2*-P522R was found to decrease the onset age of type 2 diabetes (T2D), but only in males (Fig. 4A-C).

**Figure 3.**
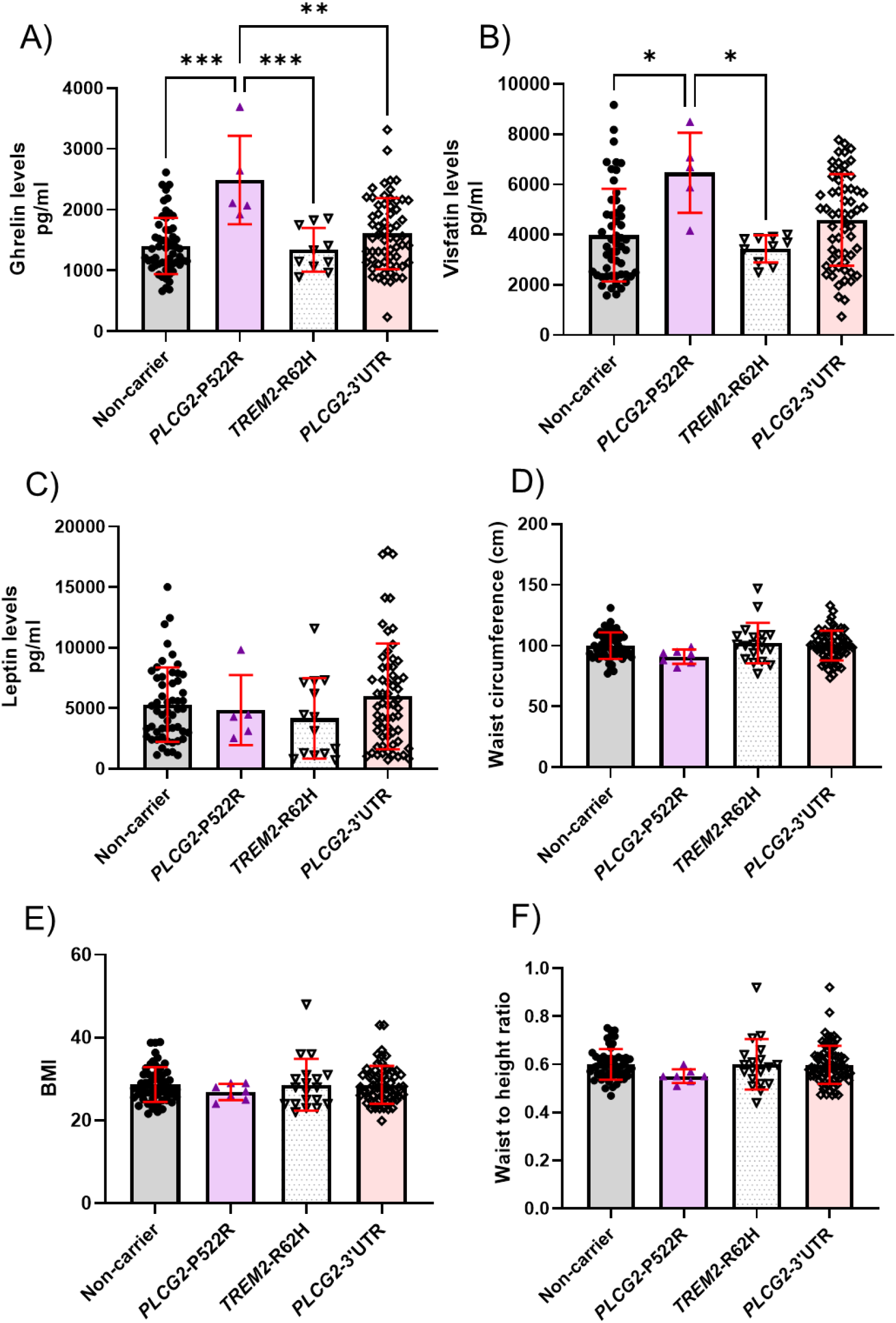
Protective *PLCG2*-P522R carriers have higher plasma ghrelin levels compared to controls, *TREM2*-R62H, and *PLCG2*-3’UTR carriers. A) Ghrelin, B) Visfatin, and C) Leptin levels in peripheral plasma samples from the FINGER cohort. D) Waist circumference, E) BMI, and F) Waist-to-height ratio of FINGER individuals. n(control)=53-56, n(P522R)=5-7, n(R62H)=10-18, and n(3’UTR)=60-63. ANOVA, Tukey’s post hoc test. Mean±SD. *<0.05, **<0.01

**Figure 4.**
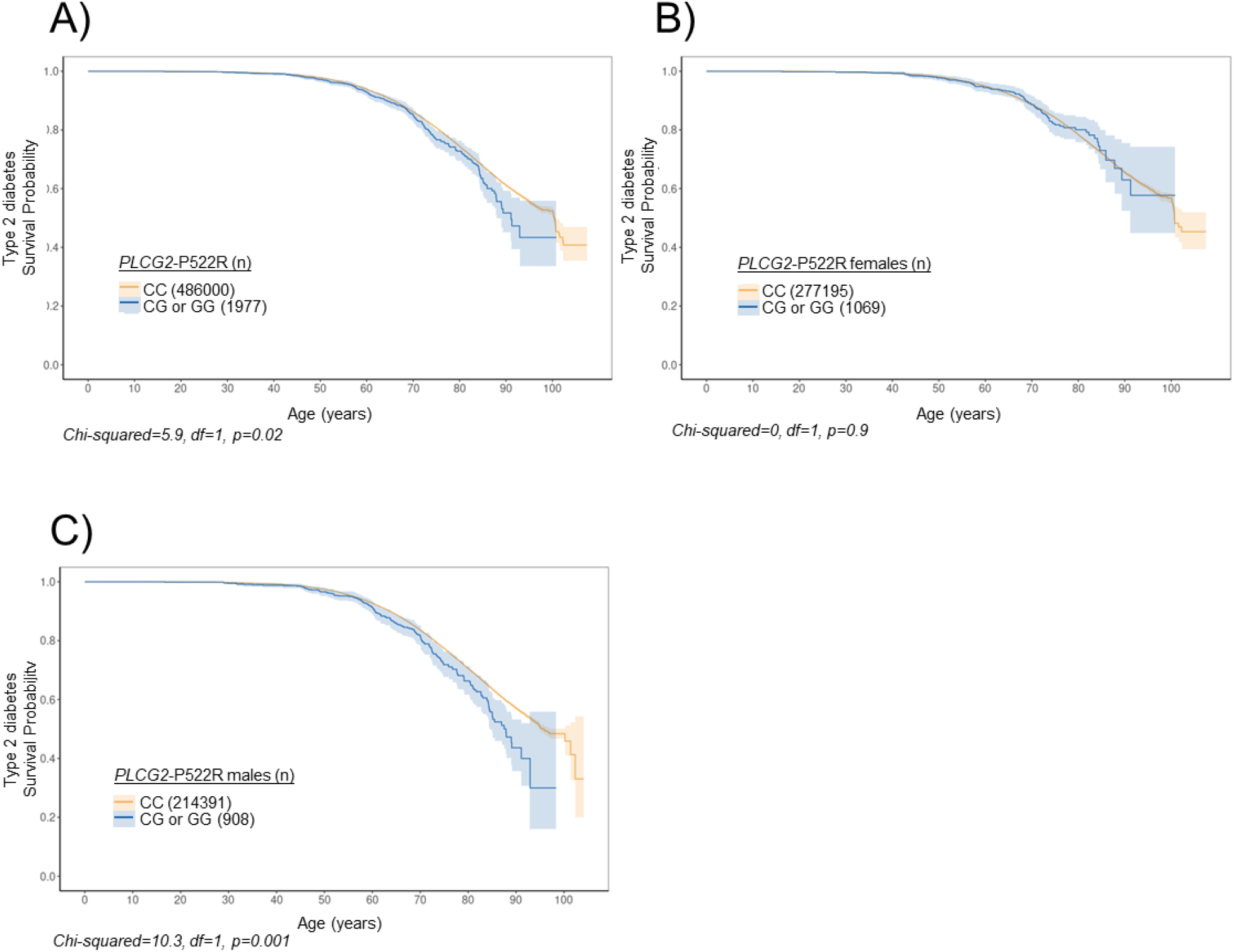
The *PLCG2*-P522R-carrying males have a higher risk of type 2 diabetes. To investigate the impact of the *PLCG2*-P522R variant on T2D onset, Kaplan-Meier curves on FinnGen endpoint data were utilized. These curves illustrate the T2D-free time in years. The x-axis indicates the age at the first diagnosis for cases and the age at the end of follow-up for the non-carrier group. Shaded area indicates 95% confidence interval. A) *PLCG2*-P522R FinnGen individuals as well as B) only females and C) only males in “T2D_WIDE” endpoint.

## 3 MATERIALS AND METHODS

This study was conducted in accordance with the ethical principles outlined in the Declaration of Helsinki.

## 3.1 Participants

FinnGen phenotype information and clinical endpoints are based on national health registries, including hospital discharge, prescription medication purchase, and cancer registers. A list of endpoints can be found from https://www.finngen.fi/en/researchers/clinical-endpoints and they can be explored at https://risteys.finngen.fi/. We have utilized information from 493,563 individuals from the FinnGen data release R12 in the analyses. In G6_ALZHEIMER endpoint, 319,783 individuals older than 50 years with *APOE* ε3/3, ε3/4, and ε4/4 genotypes were selected to investigate the effects of *APOE ε*4 on the different variant carriers on the Kaplan-Meier curves and cox analysis. In other endpoints presented in this study, all carriers of the *PLCG2*-P522R, *PLCG2*-3’UTR, and *TREM2*-R62H variants regardless of age were included. The FinnGen Study combines genome data with digital health data based on national health registers (16). FinnGen includes samples that have been collected from the Finnish biobanks as well as legacy samples, which are from previous research cohorts that has been transferred to the biobanks. The individuals in FinnGen have given written informed consent for biobank research based on the Finnish Biobank Act. Separate research cohorts that have been collected before the Finnish biobank Act (September 2013) and start of FinnGen (August 2017), have been collected based on study-specific consents and later transferred to the Finnish biobanks after approved by the Finnish Medicine Agency Fimea. LD metrics were done in the Sisu (v4.2) imputation panel used in FinnGen.

All FinnGen study subjects have undergone genome-wide genotyping. Most of the subjects have been genotyped using FinnGen ThermoFisher Axiom custom array. Approximately 70,000 subjects have been genotyped with various Illumina GWAS arrays as they originate primarily from the National Institute of Health and Welfare biobank samples that were genotyped before FinnGen. Approximately 21 million variants per individual were imputed using Finnish whole-genome reference SISu v4.2 (approximately 8,700 individuals). All genotype data is in the human genome build GRCh38/hg38. The genotype probability threshold in FinnGen was set to 0.8 for all the genotypes investigated.

The FINGER cohort characteristics (23) and study design (22) have been described previously (26). Individuals who have dementia or substantial cognitive impairment have been excluded. Here, we only utilized data from participants aged 60-78 years carrying *PLCG2-*P522R (n=7), *PLCG2*-3’UTR (n=63), or *TREM2*-R62H (n=19), and non-carriers (n=56), who did not carry any of the investigated variants. The sex distribution among non-carriers was similar to variant carriers. As *PLCG2*-3’UTR is a common variant, all the *PLCG2*-P522R and *TREM2*-R62H individuals also carry the *PLCG2*-3’UTR. Plasma biomarkers at the baseline and CRP values during one-, five-, and seven-year follow-up were used in the analysis. Forty plasma biomarkers were measured using Bioplex human diabetes 10-plex (BioRad, 10010747) assay multiplexed with adiponectin and adipsin as well as cytokine targets: eotaxin, G-CSF, GM-CSF, IFN-2Rα, IFN-γ, IL-10, IL12(p40), IL-12(p70), IL-13, IL-15, IL-1β, IL-2, IL-3, IL-4, IL5, IL-6, IL-7, IL-8, IP-10, MCP-1, MIP-1α, MIP-1β, IL-1ra, RANTES, TNF-α, TNF-β, VEGF, IL-1, and IL-17. FINGER (22) individuals were genotyped using Illumina Global Screening Assay and imputed with TOPMed reference panel as described previously by Bellenguez et al. (1).

### 3.2 FinnGen endpoints

Endpoint information can be found in the FinnGen and FinRegistery data portal Risteys (https://risteys.finngen.fi/, read 16.09.2025):

AD was defined as a diagnosis in the hospital discharge or cause of death registries with the ICD codes G30 (ICD-10) or 3310 (ICD-9). The remaining individuals were considered as controls.

T2D was defined as a diagnosis in the hospital discharge or cause of death registries with the ICD codes E11 (ICD-10) or 250.A (ICD-9). The remaining individuals were considered as controls. Individuals with pancreatitis were removed from both cases and controls.

Anxiety was defined as a diagnosis in the hospital discharge or cause of death registries with the ICD codes F41.2, F41.3, F41.8, F41.9 (ICD-10), 3000A (ICD-9), or 3000 (ICD-8). The remaining individuals, excluding those with neurotic, stress-related and somatoform disorders were considered as controls.

### 3.3 Statistical analysis

Kaplan-Meier curves were generated using R and survminer package (v 0.5.0). Also, survival analyses were performed in R using survival package (v 3.8.3). Based on the time dependence of the tested variables, cox proportional hazard model or extended cox model was used. Model fit was tested using Schoenfeld residuals. Separate *APOE* ε3/3, ε3/4, and ε4/4 groups were created to analyze the effects of *APOE ε*4 because the cox models nor the extended cox models proportional hazard assumption were met. *APOE* ε3/3 and *APOE* ε3/4 groups were analyzed using the cox model, including sex as covariate. *APOE* ε4/4 group was further divided into females and males, and the groups were then separately analyzed using the cox model. The dose-dependent effects of the variants were investigated by comparing the HRs of the different groups. Differences in BMI between genotypes were tested using Two-Way ANOVA and Tukey’s post hoc test using R (4.5.0).

Two-way ANOVA for biomarker data was performed in GraphPad Prism (v 10.4.2.633). Outliers from the data were identified using ROUT (Q=1%) and excluded from the analysis and figures. Multiple regression analyses of ghrelin, leptin, and visfatin were done in SPSS (v 29.0.0.0) adjusting for age, sex, and *APOE ε*4 carrier status. CRP levels > 10 were considered as a sign of acute inflammation and were excluded. Differences in the CRP levels over time were tested using Linear Mixed effects model using lme4 (v 1.1-37) and lmerTest (v 3.1-3) packages in R (4.4.1). Data are presented as mean ± standard deviation (SD) or standard error of the mean (SEM).

## 4 DISCUSSION

We and others have previously characterized the molecular mechanisms of the protective *PLCG2*-P522R variant in various models (6–10). Building on this, we here examined the effects of the protective *PLCG2*-P522R variant using FinnGen (16) and explored the potential plasma-based biomarkers in the FINGER cohort (22,23,26). Alongside *PLCG2*-P522R variant, we examined the newly identified common protective *PLCG2*-3’UTR variant and *TREM2*-R62H risk variant. Both *PLCG2* variants were associated with a delayed AD onset, also among *APOE ε*4 carriers, with the *PLCG2*-3’UTR variant showing a weaker, yet significant, effect. In contrast, *TREM2*-R62H variant was linked to an earlier AD onset in *APOE ε*4 carriers. Unfortunately, the low number of variant carriers limited the analysis of *PLCG2* and *TREM2* variant interaction. Furthermore, we observed increased plasma ghrelin levels in the *PLCG2*-P522R variant carriers compared to non-carriers. Altogether, the obtained results strengthen the notion that the *PLCG2* protective variants may alleviate the effects of *APOE ε*4, the strongest genetic AD risk factor and the neuroprotective effects may derive partly from increased ghrelin levels.

The protective *PLCG2*-P522R variant delayed the AD onset age in *APOE* ε3/4 carriers, but not *APOE* ε4/4 carriers, when compared to non-carriers. *APOE ε*4 is known to disrupt lipid homeostasis and immune response and to cause mitochondrial dysfunction (27–30), while *PLCG2*-P522R variant exerts opposing effects (7,9). These findings suggest that *PLCG2*-P522R variant can mitigate the *APOE ε*4-mediated risk of AD in individuals carrying one ε4 allele. *In vivo* studies have shown decreased and a more compact β-amyloid plaque area in *Plcg*2-P522R mouse brain, proposing a possible mechanism by which the *Plcg*2-P522R variant may render β-amyloid plaques less toxic to the surrounding neurons (9,10). Also, it was recently demonstrated that the *PLCG2*-P522R variant enhances immune responsiveness (31). Conversely, loss-of-function variants in *PLCG2* have been shown to impair synaptic function (5). These findings suggest that enhanced *PLCG2*-P522R-related mechanisms in both microglia and neurons may contribute to its protective effects. Although the *PLCG2*-3’UTR variant has a similar impact on the age of onset of Alzheimer’s disease as the *PLCG2*-P522R variant, the molecular mechanism underlying its protective effects remain to be elucidated.

TREM2 is one of the key receptors modulating PLCγ2 activity (12,32,33). Importantly, *TREM2* variants associate with an increased risk of AD (15,18,34–36). TREM2 deficiency leads to impaired microglial response to β-amyloid plaques, accumulation of the lipid droplets, and increased β-amyloid load in the brain (11,37). In this study we investigated whether the *TREM2*-R62H variant has opposite effects on the AD onset and biomarkers as compared to the *PLCG2*-P522R variant. We found that the *TREM2-*R62H variant decreases the onset age of AD, especially in the *APOE* ε4 carriers. Given that *APOE* ε4 is the most well know genetic risk factor for AD, it is important to understand its complex interactions with other genetic variants, including *TREM2 and PLCG2*, when considering new therapies for AD (13,14,38). In our present study, *TREM2*-R62H variant significantly increases the risk of AD only when *APOE* ε4 is also present. This differs from Thomassen et al. (38) results, which showed that *TREM2*-R62H variant significantly increases the risk of AD also in *APOE* ε3/3 carriers. This suggests that *TREM2*-R62H variant might have population specific effects. They also reported that male *APOE ε*4/4 individuals with the *TREM2*-R62H variant had higher risk of having AD as compared to females, suggesting that *TREM2*-R62H has sex-dependent effect. Given that TREM2 activation may mitigate the harmful effects of *APOE* ε4, this underscores the importance of developing novel therapies targeting components of the TREM2 pathway, such as PLCγ2, which hold significant promise as potential treatment strategies for AD. This is especially needed among individuals with increased risk of AD owing to *APOE* ε4 background, which are not fully eligible for disease-modifying treatments, such as lecanemab (39). Additionally, a deeper understanding of how sex influences AD risk across different genotypes is essential for developing effective therapeutic strategies.

Ghrelin, a neuroprotective hormone secreted by cells in the stomach, plays a key role in regulating the appetite and energy balance of the body (24,40,41). It binds to growth hormone secretagogue receptor (GHS-R), and it has been shown to protect neurons from β-amyloid-induced toxicity, reduce tau phosphorylation, and enhance synaptic plasticity (40–43). Despite its therapeutic potential, the role of ghrelin in AD pathology remains poorly understood. The *PLCG2*-P522R variant carriers showed elevated plasma levels of ghrelin and visfatin compared to non-carriers and carriers of the other investigated variants. This is an interesting finding, as elevated ghrelin levels are known to exert anti-inflammatory effects, metabolic regulation, and promote cognitive enhancement (40,42,44). Importantly, BMI or waist circumference of the *PLCG2*-P522R variant carriers did not differ from the controls or carriers of the other investigated variants, although a slight trend towards a decrease was detected in the waist circumference among the *PLCG2*-P522R-carrying females as compared to *PLCG2*-3’UTR or *TREM2*-R62H variant carriers and non-carriers. It has been shown that ghrelin can pass through the blood-brain barrier, directly affecting brain cells (43,45–47). Thus, elevated ghrelin levels in the *PLCG2*-P522R variant carriers could contribute to reduced inflammation and enhanced cognition, which are crucial in the context of neurodegeneration. In contrast, leptin levels remained unchanged in carriers of the protective *PLCG2*-P522R variant, suggesting that the hunger signal remains active. Supporting this observation, we previously demonstrated that aged knock-in mice homozygous for the *Plcg2*-P522R weighed less compared to their wild-type counterparts (9). Whether ghrelin elevation is a direct effect of *PLCG2*-P522R variant remains thus far unknown. Also, the role of visfatin is ambiguous as it has been shown to have a positive correlation with ghrelin only in metabolic syndrome or T2D (48,49). Here, we showed an earlier onset age of T2D in males but not in female *PLCG2*-P522R variant carriers. Additionally, onset of anxiety was delayed in females, suggesting sex-specific effects, consistent with prior findings (7). While therapies aimed at increasing ghrelin levels may offer benefits in neurodegeneration, it should be considered that they could also disrupt the natural metabolic balance and increase the risk of severe adverse effects.

Interestingly, ghrelin may polarize microglia to an anti-inflammatory M2 type (50) and reduce the number of inflammatory M1 type microglia in cerebral ischemic injury (51). Given this background, the protective effects of *PLCG2*-P522R variant could be linked to increased ghrelin levels, which subsequently could drive microglia polarization more towards an anti-inflammatory type. What makes this finding intriguing is that microglia lack the GHSR, the primary receptor for ghrelin (52–54). Yet, ghrelin treatment has been shown to suppress inflammatory response and reduce reactive oxygen species production in microglial cells (52,53). Furthermore, ghrelin has also been shown to act as a mitochondrional mediator and enhance mitochondrial fitness upon inflammation in macrophages (55). While the precise mechanisms underlying the effects of ghrelin on microglial signaling remain unclear, these observations suggest the involvement of GHSR–independent pathways. Hence, ghrelin could affect microglia functions indirectly by modulating other brain cell types that do express the GHSR, such as neurons (52,53,56) and astrocytes (57). Moreover, the beneficial role of ghrelin in the central nervous system has been widely discussed, including enhanced cognition (45,58– 60). However, there is still a lack of direct evidence on whether ghrelin or its active form acyl ghrelin can influence microglial metabolism or if the effects are mediated by the interaction of ghrelin with insulin signaling pathways or through neuronal pathways.

In conclusion, future studies should explore the mechanisms of how the protective *PLCG2* variants may mitigate *APOE ε*4-related impairments as it may offer new therapeutic avenues for a vast number of AD patients. Related to this, ghrelin shows an intriguing promise in modulating neurodegenerative processes, but its underlying effects, especially in *APOE ε*4 carriers, require further investigations in experimental models as well as human cohorts.

## Supporting information

FinnGen Banner_April 2025

## Data Availability

All data produced in the present study are available upon reasonable request to the authors

https://risteys.finngen.fi/

## ACKNOWLEDGEMENTS

We wish to thank the participants and investigators of the FINGER and FinnGen cohorts.

## SUPPLEMENTARY MATERIAL

FinnGen Banner updated April2025_0.xlsx

Supplementary_Figures.pdf

Supplementary_Tables.pdf

