## Supplementary Figures for "The protective *PLCG2* variants delay Alzheimer’s disease onset age in *APOE ε*4 carriers"

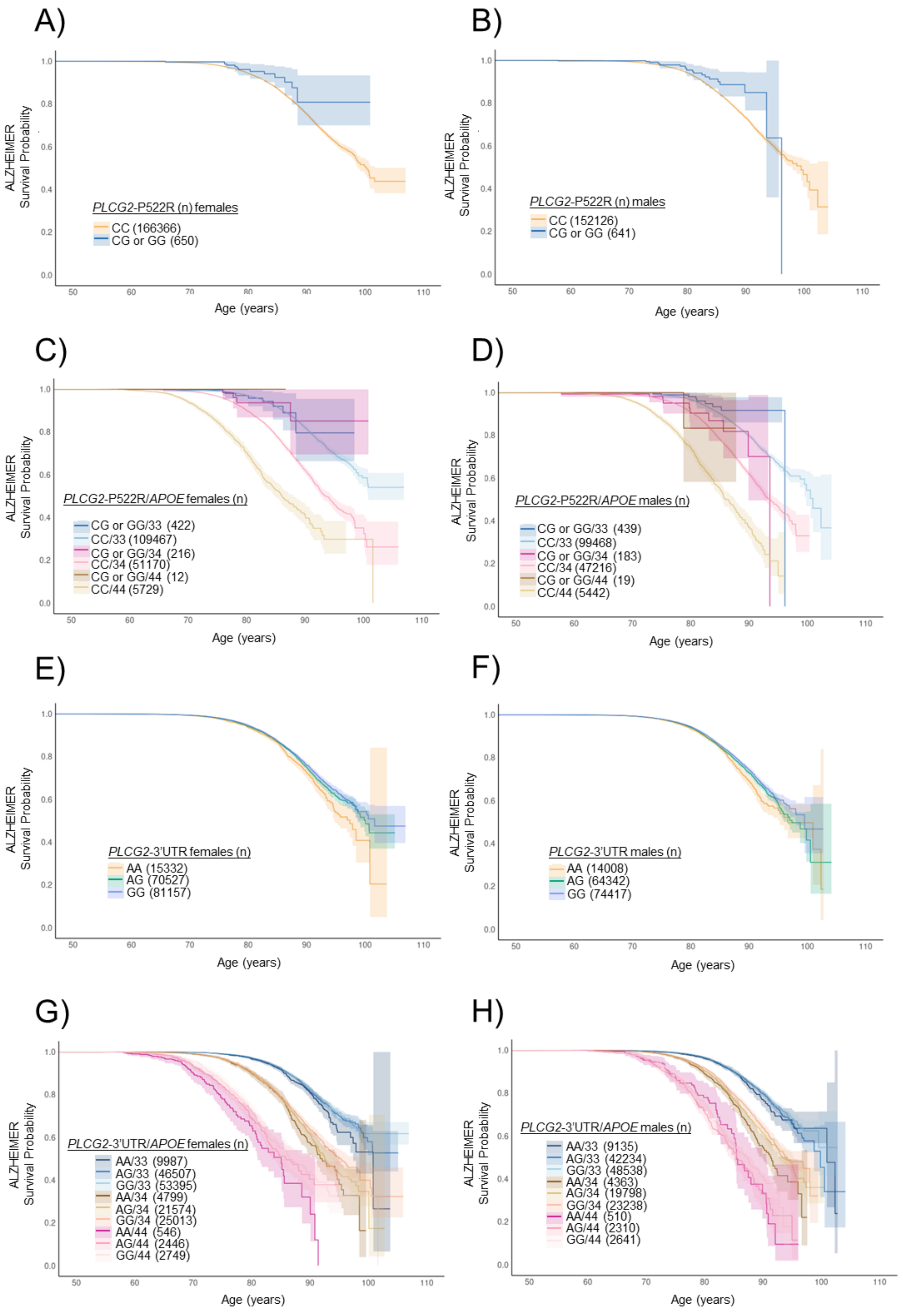
A)
ALZHEIMER
Survival Probability
Age (years)
PLCG2-P522R (n) females
CC (166366)
CG or GG (650)
B)
ALZHEIMER
Survival Probability
Age (years)
PLCG2-P522R (n) males
CC (152126)
CG or GG (641)
C)
ALZHEIMER
Survival Probability
Age (years)
PLCG2-P522R/APOE females (n)
CG or GG/33 (422)
CC/33 (109467)
CG or GG/34 (216)
CC/34 (51170)
CG or GG/44 (12)
CC/44 (5729)
D)
ALZHEIMER
Survival Probability
Age (years)
PLCG2-P522R/APOE males (n)
CG or GG/33 (439)
CC/33 (99468)
CG or GG/34 (183)
CC/34 (47216)
CG or GG/44 (19)
CC/44 (5442)
E)
ALZHEIMER
Survival Probability
Age (years)
PLCG2-3'UTR females (n)
AA (15332)
AG (70527)
GG (81157)
F)
ALZHEIMER
Survival Probability
Age (years)
PLCG2-3'UTR males (n)
AA (14008)
AG (64342)
GG (74417)
G)
ALZHEIMER
Survival Probability
Age (years)
PLCG2-3'UTR/APOE females (n)
AA/33 (9987)
AG/33 (46507)
GG/33 (53395)
AA/34 (4799)
AG/34 (21574)
GG/34 (25013)
AA/44 (546)
AG/44 (2446)
GG/44 (2749)
H)
ALZHEIMER
Survival Probability
Age (years)
PLCG2-3'UTR/APOE males (n)
AA/33 (9135)
AG/33 (42234)
GG/33 (48538)
AA/34 (4363)
AG/34 (19798)
GG/34 (23238)
AA/44 (510)
AG/44 (2310)
GG/44 (2641)

**Supplementary Figure 1. *PLCG2* variants delay AD onset age.** To investigate the impact of the variants on AD onset, Kaplan-Meier curves on FinnGen endpoint data were utilized, with the focus on *APOE* ε3- and *APOE* ε4-carrying individuals >50 years of age. The curves illustrate the AD-free time in years, starting from age 50 until AD diagnosis or the end of follow-up for the non-carrier group. Shaded area indicates 95% confidence interval. A) *PLCG2*-P522R females and B) males and as well as C) females and D) males with *APOE* ε4 allele count. E) *PLCG2*-3'UTR females and F) males as well as G) females and H) males with *APOE* ε4 allele count. *APOE*: ε3/3=33, ε3/4=34 and ε4/4=44

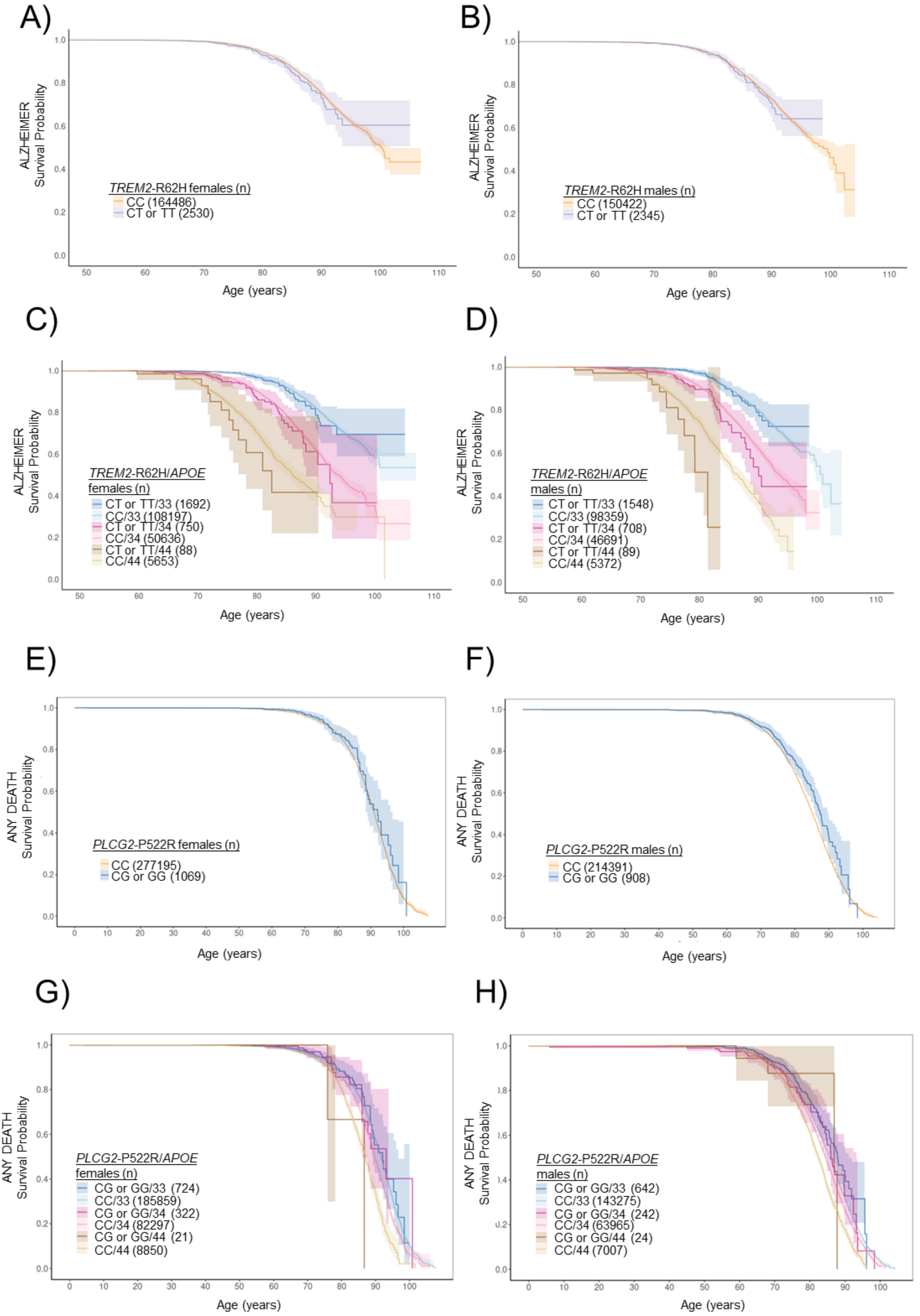
A)
ALZHEIMER
Survival Probability
TREM2-R62H females (n)
CC (164486)
CT or TT (2530)
Age (years)
B)
ALZHEIMER
Survival Probability
TREM2-R62H males (n)
CC (150422)
CT or TT (2345)
Age (years)
C)
ALZHEIMER
Survival Probability
TREM2-R62H/APOE
females (n)
CT or TT/33 (1692)
CC/33 (108197)
CT or TT/34 (750)
CC/34 (50636)
CT or TT/44 (88)
CC/44 (5653)
Age (years)
D)
ALZHEIMER
Survival Probability
TREM2-R62H/APOE
males (n)
CT or TT/33 (1548)
CC/33 (98359)
CT or TT/34 (708)
CC/34 (46691)
CT or TT/44 (89)
CC/44 (5372)
Age (years)
E)
ANY DEATH
Survival Probability
PLCG2-P522R females (n)
CC (277195)
CG or GG (1069)
Age (years)
F)
ANY DEATH
Survival Probability
PLCG2-P522R males (n)
CC (214391)
CG or GG (908)
Age (years)
G)
ANY DEATH
Survival Probability
PLCG2-P522R/APOE
females (n)
CG or GG/33 (724)
CC/33 (185859)
CG or GG/34 (322)
CC/34 (82297)
CG or GG/44 (21)
CC/44 (8850)
Age (years)
H)
ANY DEATH
Survival Probability
PLCG2-P522R/APOE
males (n)
CG or GG/33 (642)
CC/33 (143275)
CG or GG/34 (242)
CC/34 (63965)
CG or GG/44 (24)
CC/44 (7007)
Age (years)

**Supplementary Figure 2. *TREM2*-R62H decreases the onset age of *APOE ε*4 carriers.** To investigate the impact of the *TREM2*-R62H variant on AD onset, Kaplan-Meier curves on FinnGen endpoint data were utilized, with the focus on *APOE ε*3- and *APOE ε*4-carrying individuals >50 years of age. The curves illustrate the AD-free time in years, starting from age 50 until AD diagnosis or the end of follow-up for the non-carrier group. Shaded area indicates 95% confidence interval. A) *TREM2*-R62H females, B) males as well as C) females and D) males with *APOE ε*4 allele count in "ALZHEIMER" endpoint. E) *PLCG2*-P522R females and F) males as well as G) females and H) males with *APOE ε*4 allele in "ANY DEATH" endpoint. *APOE*: ε3/3=33, ε3/4=34 and ε4/4=44

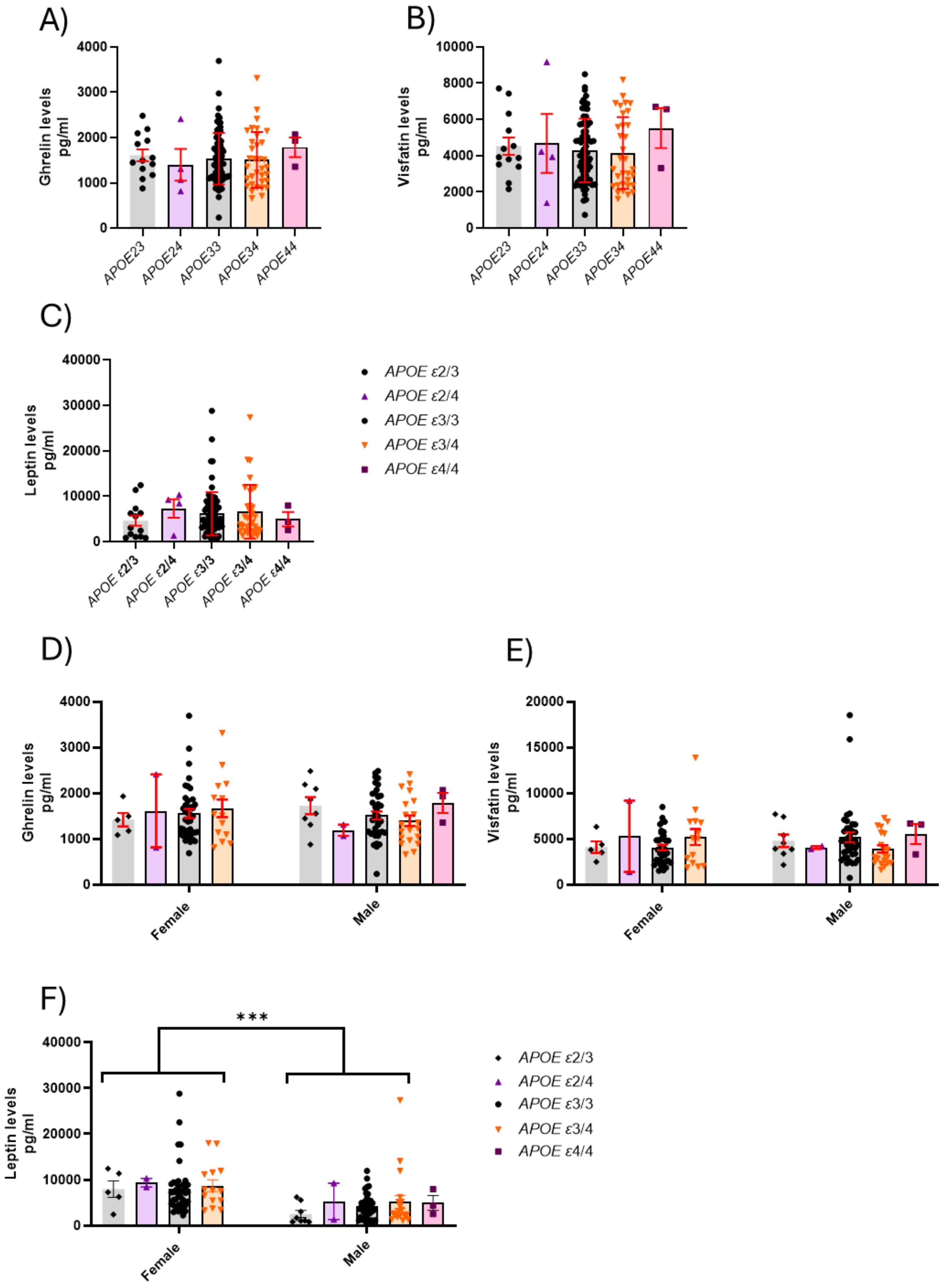
A)
Ghrelin levels pg/ml
4000
3000
2000
1000
0
APOE23
APOE24
APOE33
APOE34
APOE44
B)
Visfatin levels pg/ml
10000
8000
6000
4000
2000
0
APOE23
APOE24
APOE33
APOE34
APOE44
C)
Leptin levels pg/ml
40000
30000
20000
10000
0
APOE ε2/3
APOE ε2/4
APOE ε3/3
APOE ε3/4
APOE ε4/4
APOE ε2/3
APOE ε2/4
APOE ε3/3
APOE ε3/4
APOE ε4/4
D)
Ghrelin levels pg/ml
4000
3000
2000
1000
0
Female
Male
E)
Visfatin levels pg/ml
20000
15000
10000
5000
0
Female
Male
F)
***
Leptin levels pg/ml
40000
30000
20000
10000
0
Female
Male
APOE ε2/3
APOE ε2/4
APOE ε3/3
APOE ε3/4
APOE ε4/4

**Supplementary Figure 3. Plasma ghrelin, visfatin, and leptin levels in relation to *APOE* ε3 and *APOE* ε4 carriership.** Plasma A) ghrelin, B) visfatin, and C) leptin between *APOE* ε3 and *APOE* ε4 carriers. Plasma D) ghrelin, E) visfatin, and E) leptin levels between *APOE* ε3 and *APOE* ε4 carriers and sexes. Mean ± SEM; Independent samples t test (non-parametric) or Two-way ANOVA, Tukey's; n(*APOE* ε*2/3*)=5-13, n(*APOE* ε*2/4*)=2-4, n(*APOE* ε*3/3*)=38-77, n(*APOE* ε*3/4*)=14-34, and n(*APOE* ε*4/4*)=3

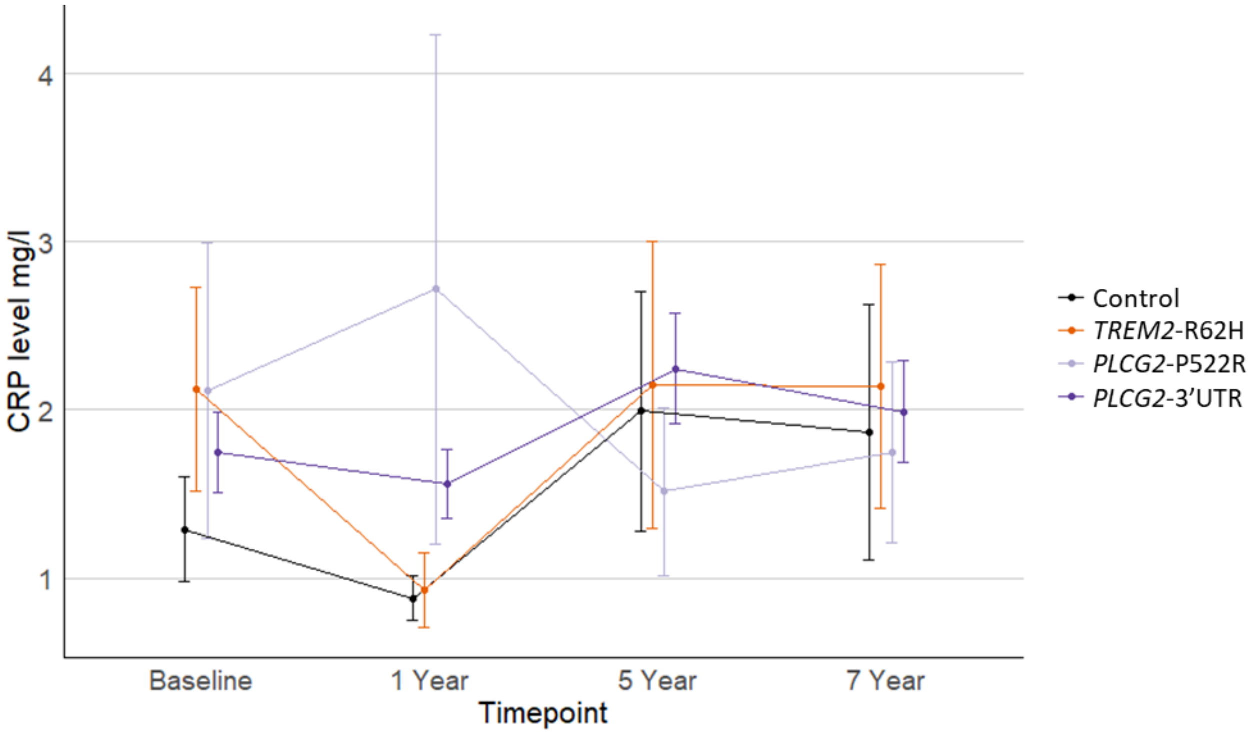


**Supplementary Figure 4. Plasma CRP levels over time in individuals from the FINGER cohort at baseline, and one-, five-, and seven-year follow-up**. Linear Mixed-Effects Model was used. Mean±SEM. n(control)=24-55, n(P522R)=4-6, n(R62H)=11-18, and n(3'UTR)=40-59.
