## Supplementary Tables for "The protective *PLCG2* variants delay Alzheimer’s disease onset age in *APOE ε*4 carriers"

**Supplementary Table 1**. The *PLCG2*-3'UTR cox regression model adjusted by sex.

| | Hazard ratio (HR) | 95% Confidence interval: lower-upper | p value |
|---|---|---|---|
| PLCG2-3'UTR heterozygous | 0.8986 | 0.8462-0.9543 | 0.000497 |
| PLCG2-3'UTR homozygous | 0.8438 | 0.7950-0.8956 | $2.35x10^{-8}$ |
| SEXmale | 1.0909 | 1.0535-1.1296 | $9.87x10^{-7}$ |

*Schoenfeld residual p=0.12*

**Supplementary table 2.** The *TREM2*-R62H cox regression model adjusted by sex.

| | Hazard ratio (HR) | 95% Confidence interval: lower-upper | p value |
|---|---|---|---|
| TREM2-R62H | 1.091 | 0.9163-1.249 | 0.204 |
| SEXmale | 1.091 | 1.0536-1.130 | $9.71x10^{-7}$ |

*Schoenfeld residual p=0.19*

**Supplementary table 3.** Comparison of plasma ghrelin levels between genotypes with age, sex, or *APOE ε4* status as co-variates in the FINGER cohort. Multilinear regression model was used.

| | *Unstandardized Coefficients* | | *Standardized Coefficients* | | | *95,0% Confidence Interval for B* | |
|---|---|---|---|---|---|---|---|
| | *B* | *Std. Error* | *Beta* | *t* | *Sig. (p)* | *Lower Bound* | *Upper Bound* |
| *(Constant)* | *3,373* | *0,291* | | *11,576* | *0,000* | *2,793* | *3,953* |
| *PLCG2* P522R seq.16.81942028.C.G | *0,227* | *0,079* | *0,312* | *2,862* | *0,005* | *0,069* | *0,385* |
| *APOE ε*4 status | *0,003* | *0,042* | *0,008* | *0,076* | *0,940* | *-0,080* | *0,086* |
| Age | *-0,003* | *0,004* | *-0,090* | *-0,815* | *0,418* | *-0,012* | *0,005* |
| Sex | *0,009* | *0,038* | *0,026* | *0,242* | *0,809* | *-0,067* | *0,086* |

*Dependent variable: logarithmic ghrelin levels*

**Supplementary table 4.** Effect of *PLCG2*-P522R variant on plasma visfatin levels between genotypes with age, sex, or *APOE ε4* status as co-variates in the FINGER cohort. Multilinear regression model was used.

| | *Unstandardized Coefficients* | | *Standardized Coefficients* | | | *95,0% Confidence Interval for B* | |
|---|---|---|---|---|---|---|---|
| | *B* | *Std. Error* | *Beta* | *t* | *Sig. (p)* | *Lower Bound* | *Upper Bound* |
| *(Constant)* | *4,021* | *0,385* | | *10,454* | *0,000* | *3,255* | *4,787* |
| *PLCG2* P522R seq.16.81942028.C.G | *0,198* | *0,105* | *0,212* | *1,894* | *0,062* | *-0,010* | *0,407* |
| *APOE ε*4status | *-0,015* | *0,055* | *-0,031* | *-0,274* | *0,785* | *-0,125* | *0,095* |
| Age | *-0,005* | *0,005* | *-0,107* | *-0,945* | *0,348* | *-0,016* | *0,006* |
| Sex | *0,009* | *0,038* | *0,026* | *0,242* | *0,809* | *-0,067* | *0,086* |

*Dependent variable: logarithmic visfatin levels*

**Supplementary 5.** Effect of *PLCG2*-P522R on plasma leptin levels between genotypes with age, sex, or *APOE ε4* status as co-variates in the FINGER cohort. Multilinear regression model was used.

| | *Unstandardized Coefficients* | | *Standardized Coefficients* | | | *95,0% Confidence Interval for B* | |
|---|---|---|---|---|---|---|---|
| | *B* | *Std. Error* | *Beta* | *t* | *Sig. (p)* | *Lower Bound* | *Upper Bound* |
| *(Constant)* | *2,714* | *0,515* | | *5,271* | *0,000* | *1,689* | *3,739* |
| *PLCG2* P522R seq.16.81942028.C.G | *-0,100* | *0,140* | *-0,065* | *-0,716* | *0,476* | *-0,380* | *0,179* |
| *APOE* ε4status | *0,110* | *0,074* | *0,135* | *1,488* | *0,141* | *-0,037* | *0,257* |
| Age | *0,004* | *0,007* | *0,045* | *0,500* | *0,618* | *-0,011* | *0,018* |
| Sex | *0,467* | *0,068* | *0,621* | *6,870* | *0,000* | *0,332* | *0,602* |

*Dependent variable: logarithmic leptin levels*
